# Predictors of Anxiety in a Sample of Preschoolers with ASD

**DOI:** 10.1101/2020.10.22.20217711

**Authors:** Samaneh Behzadpoor, Hamidreza Pouretemad, Saeed Akbari zardkhaneh

## Abstract

**Objective:** Anxiety is one of the most prevalent comorbid disorders in children with autism spectrum disorder (ASD). However, there is inconsistency in research investigating the predictors of anxiety in children with ASD. Also, most studies have focused on school-age children so research on anxiety in preschoolers with ASD has been somewhat neglected. Thus, the aim of this study was to investigate age, gender, ASD symptom severity, and functional language use as potential risk factors for developing anxiety in this sample.

**Method:** In total, 95 children with ASD and their caregivers participated in this study. The Gilliam Autism Rating Scale–Second Edition (GARS-2), and The Preschool Anxiety Scale (PAS) were used to gather data. Data were analyzed by Independent sample t-tests, MANOVA, Pearson’s r correlations, the point biserial correlation coefficient, and multiple regression analysis with the stepwise procedure.

**Results:** The findings indicated that anxiety was positively correlated with age and functional language use and negatively with ASD symptom severity. There wasn’t a significant relationship between anxiety and gender.

**Conclusion:** The findings suggest age and ASD symptom severity were significant predictors of anxiety in this sample. In other words, older children and cases with lower symptom severity are more likely to experience anxiety. It is also implied to examine the role of cognitive deficits in the development of anxiety in autism.

## Introduction

Recent research indicate that comorbid disorders are common among children with autism spectrum disorder (ASD) (1). Anxiety seems to be one of the most common psychological disorders in youth with ASD (2), but reported prevalence rates show a wide variation between 11 and 84% (3). This variation might be due to the differences in sample, methodology, assessment of anxiety, and the operationalization of anxiety across studies (4). There are some debates that cultural differences of the participants in different studies could be an explanation for the variation in reported prevalence rates of anxiety in children with ASD (5). On the basis of some research, cultural beliefs and values can protect children against or place them at risk for anxiety (6)

In order to prevent further exacerbation of comorbid anxiety in ASD, it is important to identify risk factors that predict the development of anxiety in this group. Additionally, prevention and treatment programs can also be tailored to at-risk children. Several studies identified some potential but inconsistent risk factors for developing anxiety in ASD.

Gender is one of the inconsistent but potential risk factors of anxiety in ASD. Among the studies investigated the relationship between gender and anxiety in ASD, some studies show that anxiety is more prevalent in girls than boys (5,7,8) and other studies indicated that boys with ASD have more anxiety symptoms (9). Also, there is evidence that anxiety symptoms are equally prevalent in boys and girls with ASD (10–12).

There are also mixed data regarding the prevalence rates of anxiety in ASD children with different ages. Most studies reported that anxiety in children with ASD increases with age (e.g., (9,12–14). In contrast, Wijnhoven et al. reported that anxiety was less prevalent among older than among younger children with ASD. However, some evidence showed that there is no association between age and anxiety in children with ASD (11,15).

ASD severity has also been implicated as another factor in the prediction of anxiety in children with ASD, albeit inconsistently. It has been suggested that children with less ASD symptomatology may be more vulnerable to anxiety disorders (7,12,16–18). In contrast, some research indicated that higher levels of anxiety were associated with greater ASD symptom severity (11,13,19). However, some research failed to find a relationship between ASD severity and anxiety (20,21).

It is clear that there is inconsistency in findings of research exploring the prevalence and risk factors of anxiety in children with ASD. It is important to note, while many studies have investigated anxiety in children with ASD, research specifically on anxiety in preschoolers with ASD has been somewhat overlooked (22). Research shows that there may be a relationship between the type of anxiety symptomatology and age in children with ASD (2). Understanding anxiety disorders in early childhood can inform intervention and prevention efforts to prevent later anxiety in children with ASD. Finally, Cultural differences have been frequently reported in internalized disorders including anxiety (23), but most research on anxiety in ASD has been conducted in the United States, Europe, and Australia and there are extremely few studies that investigated anxiety in Asian children with ASD (24,25). Specifically, to date, no published study has examined anxiety in Iranian children with ASD. So, the first aim of this study was to investigate the prevalence of anxiety symptoms in a group of 3- to 6-year-old Iranian children with ASD. The second aim was to examine the risk factors for anxiety in this age group of Iranian children with ASD, Specifically, gender, age, functional language use, and ASD symptom severity were investigated as potential risk factors to develop anxiety in this sample of children with ASD.

## Material and methods

### Participants

Participants consisted of 95 children with ASD (age; 37-71 months) who were referred for clinical services to two autism clinics in Tehran, Iran-Center for Treatment of Autistic Disorders (Tehran Autism) and Behara Education and Rehabilitation Center for Autistic Disorders. Inclusion criterion was sufficient knowledge of the persian language. Exclusion criteria were presence of othersevere psychological or neurological disorders and absence of parental permission. Caregivers of 95 children participated in this study (83 mothers, 8 fathers, 2 grandparents, and 2 babysitters)

### Study procedure

A total of 95 caregivers of children with ASD who interested in participating in the study completed a questionnaire on anxiety (Preschool Anxiety Scale (PAS)) and ASD severity (Gilliam Autism Rating Scale–Second Edition). First, parents received some information about the stud by phone. If parents were interested in participation, they filled the mentioned questionnaires.Both questionnaires completed through an interview with caregivers by a trained psychologist. Caregivers also completed a Demographic Information form designed by authors.

### Measures

The Gilliam Autism Rating Scale–Second Edition (GARS-2): it is a tool for the assessment and diagnosis of autism based on DSM-IV for individuals between the ages of 3 and 22 (26). GARS- 2 gathers information in three areas (Stereotyped Behaviors, Communication, and Social Interaction) and it also contains a developmental history. It can give us a standardized score of autism severity. This is a standardized scale, which include 42 items divided into three subscales based on the definition of autism on the diagnostic criteria for autistic disorder presnted in DSM- IV-TR. It is validated and standardized on Iranian population by Samadi, and McConkey (27). The Preschool Anxiety Scale (PAS): It is developed by Spence (28) to evaluate a wide range of anxiety symptoms in preschoolers aged 31 to 83 months based on Diagnosis and Statistical Manual of Mental Disorders-4th ed (DSM–IV) and consists of 28 items into five subscales: generalized anxiety disorder, social anxiety disorder or social phobia, separation anxiety disorder, obsessive-compulsive disorder, and physical injury fears. The PAS also provides a total score of anxiety. Ghanbari et al (2011) in their research on irainian children showed that ubscales of PAS had moderate to high internal consistency and good reliabiity. Validity evaluations also yielded positive results (29)

The PAS has previously been employed and indicated to be a valid measure of anxiety levels in preschoolers with ASD (30,31). The score of obsessive-compulsive subscale was not included, because children with ASD serve this behavior for self-soothing or regulation of emotions (32) and it is not related to anxiety. Furthermore, in the DSM5, the obsessive-compulsive disorder is considered as an anxiety disorder no more (33). Moreover, like Wijnhoven et al., we also defined the physical injury fears as “specific phobia” in this study, because the items of the PAS in this subscale do not only include physical injury fears but also include other specific fears such as fear of darkness and fear of heights (5).

### Analysis

Independent sample t-test and MANOVA were used to examine gender differences. Pearson’s r correlations between PAZ total anxiety, age, and ASD severity were calculated. The point-biserial correlation coefficient is used to examine the relation between the level of total anxiety and gender. Because of inconsistencies in literature about predictors of anxiety in ASD, Multiple regression analysis with the stepwise procedure was used to reveal contributions of the dichotomous variables, gender (Male=0; Female=1) and functional language use (Yes=1; No=0), and scores on two continuous variables including age and autism symptom severity. Data were statistically analyzed by SPSS software (Version 16).

## Results

### Socio-demographic characteristics

The demographic information of children and their caregivers who participated in this study are shown in table 1. As shown in Table 1, 72 children were boys, and 23 of them were girls. All children were diagnosed with autism. Among children, 43.2% were between 3-4 years, 43.2% were between 4-5 years, and 13.6% were 5-6 years old. The majority of caregivers were college-educated.

**Table 1.**
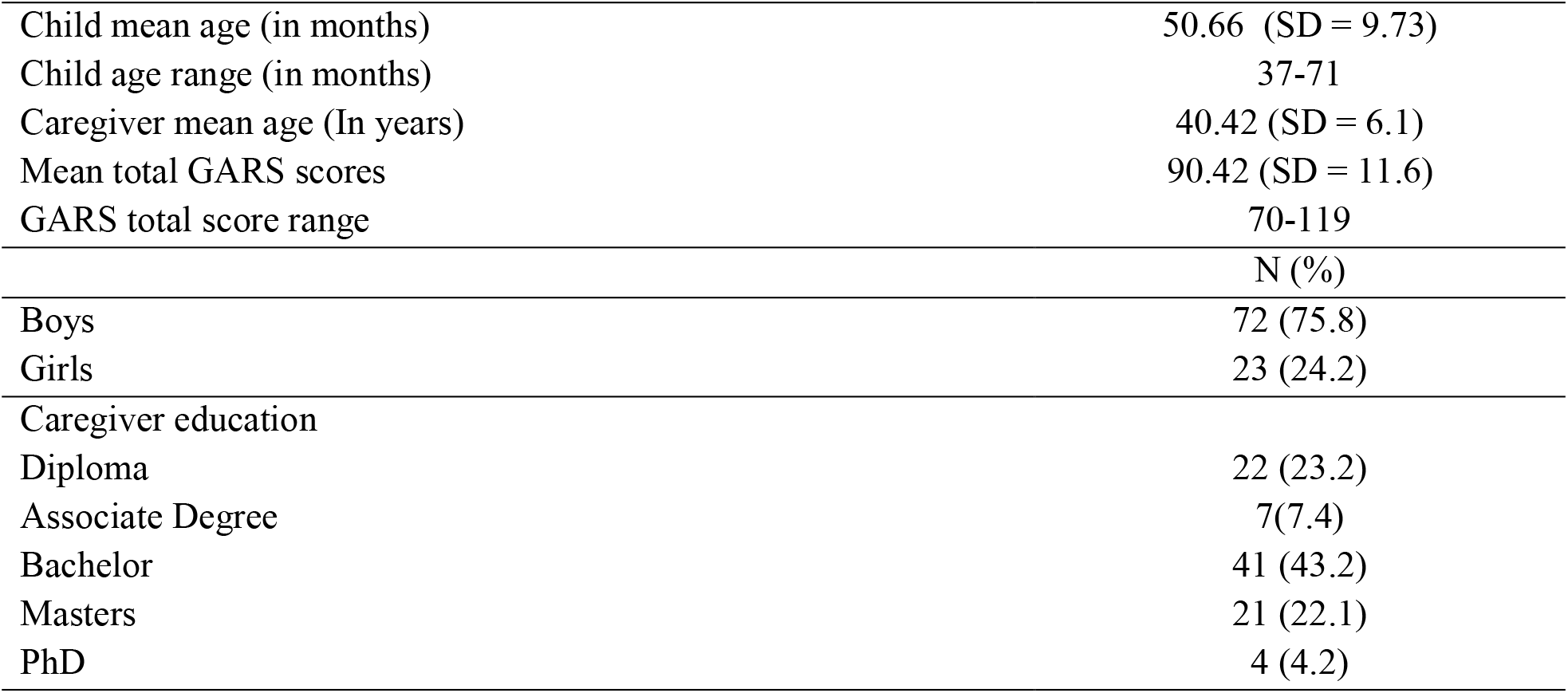
Demographic data for the children and their caregivers

|  |  |
| --- | --- |
| Child mean age (in months) | 50.66 (SD = 9.73) |
| Child age range (in months) | 37-71 |
| Caregiver mean age (In years) | 40.42 (SD = 6.1) |
| Mean total GARS scores | 90.42 (SD = 11.6) |
| GARS total score range | 70-119 |
|  | N (%) |
| Boys | 72 (75.8) |
| Girls | 23 (24.2) |
| Caregiver education |  |
| Diploma | 22 (23.2) |
| Associate Degree | 7(7.4) |
| Bachelor | 41 (43.2) |
| Masters | 21 (22.1) |
| PhD | 4 (4.2) |

### Gender differences in total anxiety and anxiety symptoms

We performed an independent t-test and MANOVA to examine the gender differences in total anxiety and anxiety symptoms among children with ASD. Tables 2, shows the mean and standard deviation of total anxiety in boys and girl and results of t-test. The results indicated that there is no significant differences in total anxiety between girls and boys. Also, the results of MANOVA showed that there are no significant differences between girls and boys in anxiety symptoms.

**Table 2.** Gender differences in total anxiety

|  | Male<br>(n = 72)<br>Mean(S.D) | Female<br>(n = 23)<br>Mean(S.D) | t | p |
| --- | --- | --- | --- | --- |
| Total anxiety | 27.69 (14.03) | 24.43 (14.91) | 0.955 | 0.342 |

### Relationship between risk factors and anxiety

The correlations between the four risk factors examined in this study (age, gender, functional language use, and autism symptom severity) and total anxiety were examined (Table 4). As shown in table 4, There were no significant relationships between gender and overall anxiety (r = −0.099, p > 0.05). Total anxiety score was positively correlated with age (r = 0.33, p < 0.01) and functional language use (r = 0.319, p < 0.01). Finally, there was a significant negative correlation between total anxiety and autism symptom severity (r = −0.354, p < 0.01).

**Table 3.**
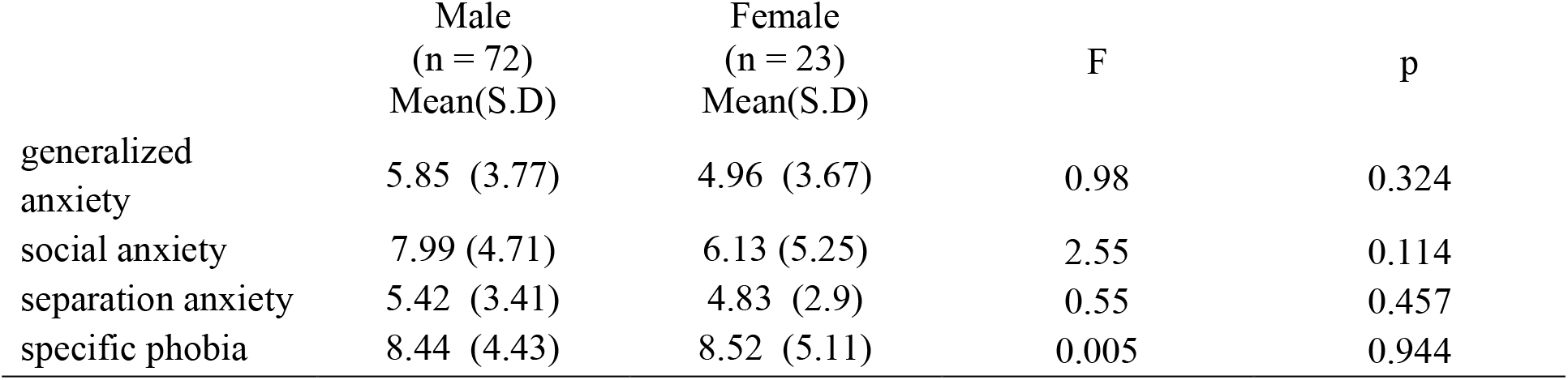
Gender differences in anxiety symptoms

|  | Male<br>(n = 72)<br>Mean(S.D) | Female<br>(n = 23)<br>Mean(S.D) | F | p |
| --- | --- | --- | --- | --- |
| generalized anxiety | 5.85 (3.77) | 4.96 (3.67) | 0.98 | 0.324 |
| social anxiety | 7.99 (4.71) | 6.13 (5.25) | 2.55 | 0.114 |
| separation anxiety | 5.42 (3.41) | 4.83 (2.9) | 0.55 | 0.457 |
| specific phobia | 8.44 (4.43) | 8.52 (5.11) | 0.005 | 0.944 |

**Table 4.** Correlation between scores of total anxiety and gender, functional language use, autism symptom severity, and age

|  | Gender | functional language use | Autism symptom severity | age |
| --- | --- | --- | --- | --- |
| Total anxiety | -.099 | .318** | -.354** | 0.330** |

\*\*. Correlation is significant at the 0.01 level

### Prediction of anxiety based on risk factors

Stepwise regression analysis was used to examine the role of independent variables in the prediction of total anxiety. The results presented in Tables 5 and 6. The correlation coefficients showed that there was no significant relationship between gender and anxiety (see table 4), so gender didn’t include as a predictor variable into the regression model. By including three variables, age, autism symptom severity, and functional language use into the stepwise linear regression analysis, only the autism symptom severity and age were significant predictors of anxiety (P<0.01). Functional language use was excluded variable and did not have a significant role in predicting total anxiety (P>.05). The model explained 22.7 of the total variance in predicting self-harm (F = 12.153, p < 0.01). symptom severity was the strongest predictor and entered the model first, followed by age. The first predictor accounted for 12.5% of the variance in anxiety (F = 13.324, P < 0.01). Age emerged as the second most potent predictor of anxiety and accounted for 10.2% of the variance in anxiety (see table 5). Table 6 shows that total anxiety decreases with symptom severity but increases with age.

**Table 5.**
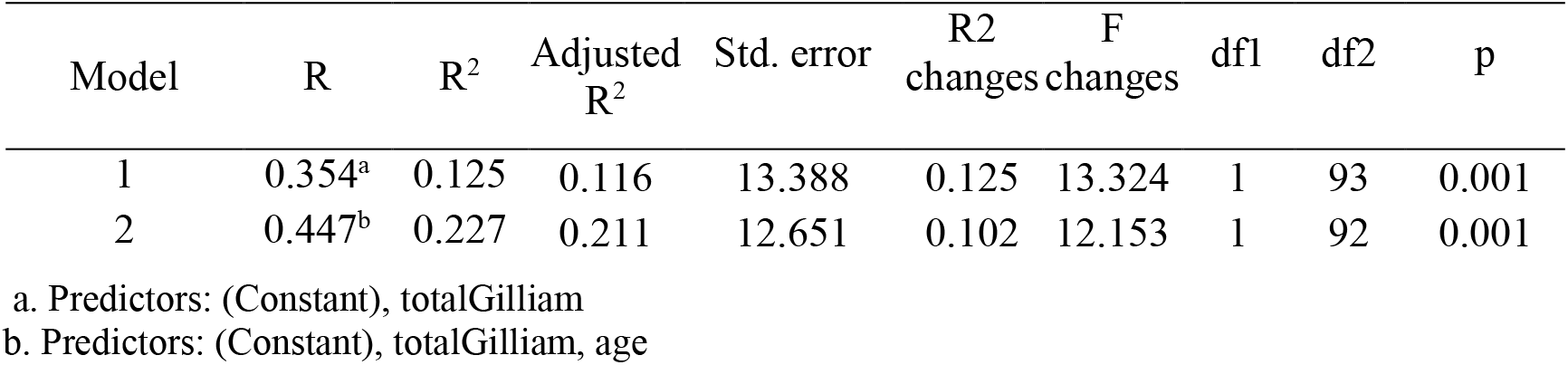
Summary of the Regression Model for the Prediction of anxiety

| Model | R | R<sup>2</sup> | Adjusted R<sup>2</sup> | Std. error | R2 changes | F changes | df1 | df2 | p |
| --- | --- | --- | --- | --- | --- | --- | --- | --- | --- |
| 1 | 0.354<sup>a</sup> | 0.125 | 0.116 | 13.388 | 0.125 | 13.324 | 1 | 93 | 0.001 |
| 2 | 0.447<sup>b</sup> | 0.227 | 0.211 | 12.651 | 0.102 | 12.153 | 1 | 92 | 0.001 |

a. Predictors: (Constant), totalGilliam
b. Predictors: (Constant), totalGilliam, age

**Table 6.**
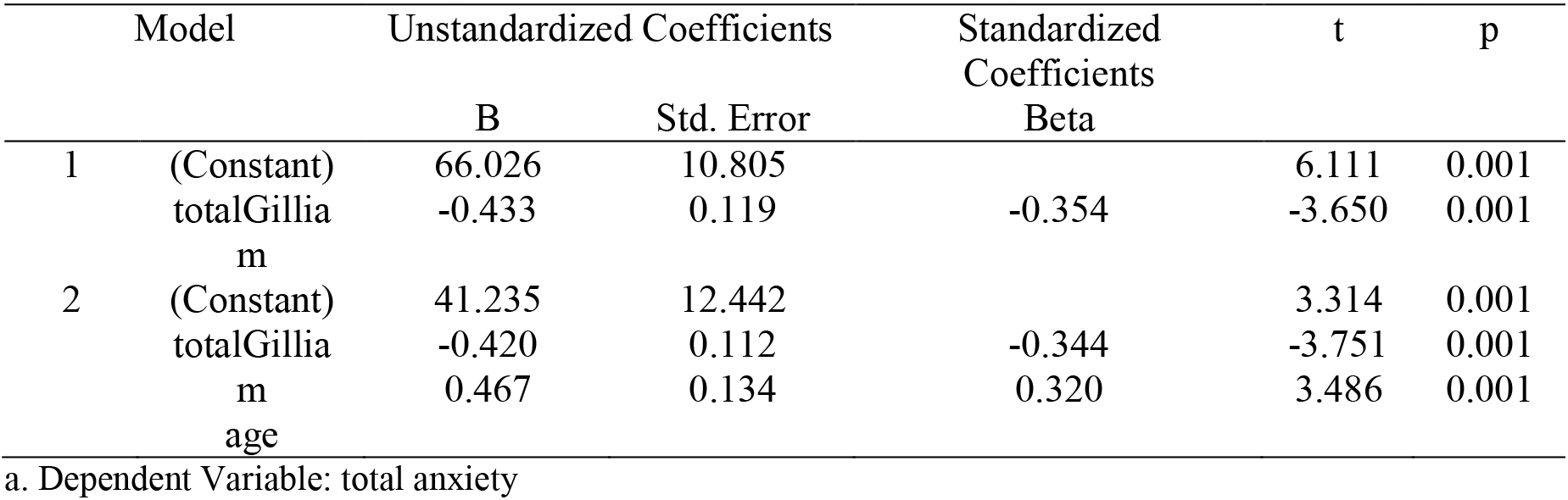
Coefficients^a^ of Regression for the Prediction of anxiety

## Discussion

The first purpose of this study was to present an overview of the prevalence of anxiety symptoms in an Iranian clinical sample of children with ASD. The findings showed that 57.9% of the children had at least one clinical anxiety symptom according to caregivers. The caregiver-rated prevalence of anxiety (57.9%) found in this study was approximately similar to the prevalence rates in some research such as the study of Strang et al. and Vasa et al. But in the research of Wijnhoven et al., more than 60% of the ASD children according to children and more than 80% of the children according to parents had at least subclinical anxiety symptoms. However, other studies reported that only 22 (34) and 41.9% (21) of the children with ASD had anxiety symptoms. Differences in the cultural background of the participants and in tools used to assess anxiety symptoms may partly explain the varying rate of anxiety symptoms reported in studies. Finally, anxiety levels may differ as a result of the differences in the age group of the sample.

The second aim was to investigate the risk factors for anxiety in this sample of children with ASD. More specifically, age, gender, ASD symptom severity, and functional language use were investigated as risk factors for anxiety in children with ASD. Regarding the differential prevalence rates of anxiety in boys and girls with ASD, no gender differences have been identified and the results showed that there weren’t any relations between gender and anxiety. This finding was in line with research showing that there are no gender differences in anxiety between girls and boys with ASD (10–12). However, this finding contradicts previous research that has shown girls with ASD are more anxious than boys (8,35,36) or boys experience more anxiety than girls (9). This result might be due to the unequal ratio of boys and girls in the sample group or differences of age and symptom severity between girls and boys who participated in this study that can influence on anxiety level.

Additionally, Our results indicated that total anxiety symptom was more prevalent among older than among younger children with ASD and age was a predictor of anxiety. This finding is in line with previous studies that have shown that anxiety increases with age in the ASD population (e.g., 8,11–13) and is inconsistent with research citing a decrease in anxiety with age in youth with ASD (e.g., 33). Furthermore, the finding that ASD children with a low ASD symptom severity had higher anxiety levels is consistent with research showing that more severe ASD symptoms are related to fewer anxiety levels (12,16–18,35), but is inconsistent with the finding showing that children with greater ASD symptoms are more vulnerable to clinical anxiety symptoms (11,13,19). This finding also extends the previous results by showing that ASD symptom severity in a multivariate model is the strongest predictor of anxiety in ASD as compared to other factors such as age and functional language use. We can explain these results according to cognitive development. Young children are growing cognitively and their cognitive abilities increase with age. Theories of emotional development discuss that cognitive development plays an important role in emotions including anxiety (e.g., 34,35). Some researchers found a positive relationship between cognitive development and the level of anxiety (39–41). Research on ASD children also found a positive relationship between cognitive ability and the level of anxiety (13,42). Younger children and children with more severe ASD symptoms may have more problems in expressing their emotions including anxiety because of their less communication and cognitive skills. Mayes states that anxiety symptoms likely require self-perception, levels of cognition, and social awareness. These requirements are not present in very young children with autism and severe cases of ASD, so they may be less aware of their impairments and their effects (13). In this line, Mazurek and Kanne believe that weak emotional understanding and perspective-taking skills may prevent the development of anxiety symptoms in children with more severe ASD symptoms. According to these explanations, experiencing anxiety requires some cognitive abilities impaired in children with severe ASD symptoms. So it is likely that children with more severe symptoms that have more cognitive impairments can’t experience anxiety and some anxiety symptoms reported in these children may be characteristics of ASD or symptoms of fear.

In addition, the present study revealed that higher functional language use was associated with some higher levels of anxiety. But this variable couldn’t predict anxiety significantly. This finding was in line with research showing that higher levels of anxiety were associated with the presence of functional language use (11). One explanation is that children with lower functional language may have difficulty expressing their feelings. Another explanation is that language and cognitive development are interrelated. Language development research recognized that language processing is cognition and language use is distributed cognition (43). Again, this may be explained by relatively low cognitive levels in ASD children with lower levels of functional language acting as a buffer against developing anxiety symptoms.

## Limitation

The findings of the present study are promising, but there are some limitations in this study that should be considered. It is possible that impairments related to the core ASD symptoms (e.g., social withdrawal) not pure anxiety symptoms were measured with the PAZ. As a consequence, the caregivers of ASD children may not have accurately reported the anxiety symptoms in ASD children. Also, this study relied on caregiver report and caregiver perceptions of anxiety symptoms may differ from those of their child. Furthermore, caregiver characteristics such as caregiver anxiety may influence on caregiver report on children’s anxiety symptoms (44). Despite these potential limitations of the caregiver-rating of anxiety, the study of Sukhodolsky et al. suggest that caregiver ratings could be a useful source of information about anxiety symptoms in children with ASD. Another limitation is that lack of psychometric characteristics for the ASD preschoolers of the PAZ may lead to an unreliable evaluation of anxiety. However, this scale has previously been used to measure anxiety in preschoolers with ASD and indicated to be a valid measure of anxiety levels in this population (30,31).

## Conclusion

This study was the first research examined anxiety in Iranian children with ASD and provided preliminary findings regarding prevalence and predictors of anxiety reported by caregivers. It is evident that 57.9% of the participating preschoolers with ASD had at least one clinical anxiety symptoms. No gender differences in any of the anxiety symptoms were found in this study. Age and functional language use were positively related to anxiety and ASD symptom severity was inversely correlated with anxiety. Among these variables, ASD symptom severity and age can significantly predict anxiety and ASD symptom severity was the strongest predictor of anxiety in this model. In light of current findings and contradictory findings in past research, it is clear that further investigation is needed to identify additional predictors of anxiety and the role of the combination of different variables in explanation of anxiety in preschoolers with ASD. It is also suggested to examine the role of cognitive deficits in the development of anxiety in autism. Children with ASD, specifically children with more severe symptoms, have fundamental cognitive deficits that can influence their experienceing and expressing anxiety. Therefore, in order to provide an appropriate explanation for anxiety in children with ASD, more research is needed o determine which cognitive impairments in these individuals are related to anxiety or may influence the development of anxiety in this population. Thus, an important future direction is to provide a conceptualization of anxiety in children with autism regarding their cognitive deficits.

## Data Availability

The data that support the findings of this study are available from the corresponding author, upon reasonable request.

## Acknowledgement

The authors are grateful to the children with ASD and their caregivers for participating in this research study. We also thank the Center for Treatment of Autistic Disorders (Tehran Autism) and Behara Education and Rehabilitation Center for Autistic Disorders for their support of this study.

## Conflict of Interest

The authors declared no potential conflicts of interest with respect to the research, authorship, and/or publication of this article.

## Notes

### Competing Interest Statement

The authors have declared no competing interest.

### Funding Statement

The authors received no financial support for the research, writing, and/or publication of this article.

### Author Declarations

All procedures were approved by the ethical committee of Shahid Beheshti University (ethical code: IR.SBU.REC.1398.023).

## References

1. Salazar F, Baird G, Chandler S, Tseng E, O’sullivan T, Howlin P, et al. Co-occurring Psychiatric Disorders in Preschool and Elementary School-Aged Children with Autism Spectrum Disorder. J Autism Dev Disord. 2015 Aug 27;45(8):2283–94.

2. van Steensel FJA, Bögels SM, Perrin S. Anxiety Disorders in Children and Adolescents with Autistic Spectrum Disorders: A Meta-Analysis. Clin Child Fam Psychol Rev [Internet]. 2011 Sep 7 [cited 2019 Feb 20];14(3):302–17. Available from: http://www.ncbi.nlm.nih.gov/pubmed/21735077

3. White SW, Oswald D, Ollendick T, Scahill L. Anxiety in children and adolescents with autism spectrum disorders. Clin Psychol Rev [Internet]. 2009 Apr [cited 2019 Apr 15];29(3):216–29. Available from: http://www.ncbi.nlm.nih.gov/pubmed/19223098

4. Kerns CM, Kendall PC. The Presentation and Classification of Anxiety in Autism Spectrum Disorder. Clin Psychol Sci Pract [Internet]. 2012 Dec 1 [cited 2019 Jul 14];19(4):323–47. Available from: http://doi.wiley.com/10.1111/cpsp.12009

5. Wijnhoven LAMW, Creemers DHM, Vermulst AA, Granic I. Prevalence and Risk Factors of Anxiety in a Clinical Dutch Sample of Children with an Autism Spectrum Disorder. Front psychiatry [Internet]. 2018 [cited 2019 Apr 8];9(50):1–10. Available from: http://www.ncbi.nlm.nih.gov/pubmed/29551982

6. Varela EE, Hensley-Maloney L. The influence of culture on anxiety in latino youth: A review [Internet]. Vol. 12, Clinical Child and Family Psychology Review. Clin Child Fam Psychol Rev; 2009 [cited 2020 Oct 20]. p. 217–33. Available from: https://pubmed.ncbi.nlm.nih.gov/19277865/

7. Gadow KD, Devincent CJ, Pomeroy J, Azizian A. Comparison of DSM-IV symptoms in elementary school-age children with PDD versus clinic and community samples. Autism. 2005;9(4):392–415.

8. May T, Cornish K, Rinehart N. Does gender matter? A one year follow-up of autistic, attention and anxiety symptoms in high-functioning children with autism spectrum disorder. J Autism Dev Disord [Internet]. 2014;44(5):1077–86. Available from: https://www.ncbi.nlm.nih.gov/pubmed/24105364

9. Dubin AH, Lieberman-Betz, R., Michele Lease A. Investigation of Individual Factors Associated with Anxiety in Youth with Autism Spectrum Disorders. J Autism Dev Disord [Internet]. 2015;45(9):2947–60. Available from: https://www.ncbi.nlm.nih.gov/pubmed/25917383

10. Magiati I, Ong C, Lim XY, Tan JW-L, Ong AYL, Patrycia F, et al. Anxiety symptoms in young people with autism spectrum disorder attending special schools: Associations with gender, adaptive functioning and autism symptomatology. Autism [Internet]. 2016 Apr 27 [cited 2019 Feb 20];20(3):306–20. Available from: http://www.ncbi.nlm.nih.gov/pubmed/25916865

11. Sukhodolsky DG, Scahill L, Gadow KD, Arnold LE, Aman MG, McDougle CJ, et al. Parent-Rated Anxiety Symptoms in Children with Pervasive Developmental Disorders: Frequency and Association with Core Autism Symptoms and Cognitive Functioning. J Abnorm Child Psychol [Internet]. 2008 Jan 3 [cited 2019 Feb 20];36(1):117–28. Available from: http://www.ncbi.nlm.nih.gov/pubmed/17674186

12. Vasa RA, Kalb L, Mazurek M, Kanne S, Freedman B, Keefer A, et al. Age-related differences in the prevalence and correlates of anxiety in youth with autism spectrum disorders. Res Autism Spectr Disord [Internet]. 2013 Nov 1 [cited 2019 Apr 8];7(11):1358–69. Available from: https://www.sciencedirect.com/science/article/pii/S1750946713001220

13. Mayes SD, Calhoun SL, Murray MJ, Zahid J. Variables Associated with Anxiety and Depression in Children with Autism. J Dev Phys Disabil [Internet]. 2011 Aug 23 [cited 2019 Jul 10];23(4):325–37. Available from: http://link.springer.com/10.1007/s10882-011-9231-7

14. Williams S, Leader G, Mannion A, Chen J. An investigation of anxiety in children and adolescents with autism spectrum disorder. Res Autism Spectr Disord [Internet]. 2015;10:30–40. Available from: https://www.sciencedirect.com/science/article/pii/S175094671400258X

15. Strang JF, Kenworthy L, Daniolos P, Case L, Wills MC, Martin A, et al. Depression and anxiety symptoms in children and adolescents with autism spectrum disorders without intellectual disability. Res Autism Spectr Disord [Internet]. 2012;6(1):406–12. Available from: https://www.ncbi.nlm.nih.gov/pmc/articles/PMC3355529/

16. Eussen MLJM, Van Gool AR, Verheij F, De Nijs PFA, Verhulst FC, Greaves-Lord K. The association of quality of social relations, symptom severity and intelligence with anxiety in children with autism spectrum disorders. Autism [Internet]. 2013 Nov [cited 2019 Feb 20];17(6):723–35. Available from: http://www.ncbi.nlm.nih.gov/pubmed/22917843

17. Mazurek MO, Kanne SM. Friendship and Internalizing Symptoms Among Children and Adolescents with ASD. J Autism Dev Disord [Internet]. 2010 Dec 20 [cited 2019 Apr 15];40(12):1512–20. Available from: http://www.ncbi.nlm.nih.gov/pubmed/20405193

18. Snow A V., Lecavalier L. Comparing Autism, PDD-NOS, and Other Developmental Disabilities on Parent-Reported Behavior Problems: Little Evidence for ASD Subtype Validity. J Autism Dev Disord [Internet]. 2011 Mar 16 [cited 2019 Apr 15];41(3):302–10. Available from: http://www.ncbi.nlm.nih.gov/pubmed/20556500

19. Rosenberg RE, Kaufmann WE, Law JK, Law PA. Parent Report of Community Psychiatric Comorbid Diagnoses in Autism Spectrum Disorders. Autism Res Treat [Internet]. 2011;2011:1–10. Available from: https://www.hindawi.com/journals/aurt/2011/405849/

20. Renno P, Wood JJ. Discriminant and convergent validity of the anxiety construct in children with autism spectrum disorders. J Autism Dev Disord. 2013 Sep;43(9):2135–46.

21. Simonoff E, Pickles A, Charman T, Chandler S, Loucas T, Baird G. Psychiatric Disorders in Children With Autism Spectrum Disorders: Prevalence, Comorbidity, and Associated Factors in a Population-Derived Sample. J Am Acad Child Adolesc Psychiatry [Internet]. 2008 Aug [cited 2019 Jul 21];47(8):921–9. Available from: https://www.ncbi.nlm.nih.gov/pubmed/18645422

22. Keen D, Adams D, Simpson K, den Houting J, Roberts J. Anxiety-related symptomatology in young children on the autism spectrum. Autism [Internet]. 2019 [cited 2020 Jan 14];23(2):350–8. Available from: http://www.ncbi.nlm.nih.gov/pubmed/29202607

23. Anderson ER, Mayes LC. Race/ethnicity and internalizing disorders in youth: A review. Vol. 30, Clinical Psychology Review. 2010. p. 338–48.

24. Magiati I, Ozsivadjian A. Phenomenology and Presentation of Anxiety in Autism Spectrum Disorder. In: Connor M. Kerns, Patricia Renno, Eric A. Storch, Philip C. Kendall, Jeffrey J. Wood, editors. Anxiety in Children and Adolescents with Autism Spectrum Disorder [Internet]. Academic Press; 2017 [cited 2019 Jul 8]. p. 33–54. Available from: https://www.sciencedirect.com/science/article/pii/B978012805122100003X

25. Ooi YP, Rescorla L, Sung M, Fung DSS, Woo B, Ang RP. Comparisons between autism spectrum disorders and anxiety disorders: Findings from a clinic sample in Singapore. Asia-Pacific Psychiatry. 2014 Mar;6(1):46–53.

26. Montgomery JM, Newton B, Smith C. Test Review: Gilliam, J. (2006). GARS-2: Gilliam Autism Rating Scale—Second Edition. Austin, TX: PRO-ED. J Psychoeduc Assess [Internet]. 2008 Dec 9 [cited 2020 Jan 21];26(4):395–401. Available from: http://journals.sagepub.com/doi/10.1177/0734282908317116

27. Samadi SA, McConkey R. The utility of the Gilliam autism rating scale for identifying Iranian children with autism. Disabil Rehabil [Internet]. 2014 [cited 2020 Oct 20];36(6):452–6. Available from: https://pubmed.ncbi.nlm.nih.gov/23738615/

28. Spence SH, Rapee R, McDonald C, Ingram M. The structure of anxiety symptoms among preschoolers. Behav Res Ther [Internet]. 2001 Nov [cited 2019 Aug 7];39(11):1293–316. Available from: http://www.ncbi.nlm.nih.gov/pubmed/11686265

29. Ghanbari, Saeed., Khanmohamadi, Maryam., Khodapanahi, Mohammad Karim., Mazaheri, Mohammad Ali., Gholamali Lavasani M. Study of Psychometric Properties of Preschool Anxiety Scale (PAS). J Psychol. 2011;15(3):222–34.

30. MacLennan K, Roach L, Tavassoli T. The Relationship Between Sensory Reactivity Differences and Anxiety Subtypes in Autistic Children. Autism Res [Internet]. 2020 Jan 7 [cited 2020 Jan 21];aur.2259. Available from: https://onlinelibrary.wiley.com/doi/abs/10.1002/aur.2259

31. Potter LA, Scholze DA, Biag HMB, Schneider A, Chen Y, Nguyen D V., et al. A Randomized Controlled Trial of Sertraline in Young Children With Autism Spectrum Disorder. Front Psychiatry [Internet]. 2019 Nov 6 [cited 2020 Jan 21];10. Available from: https://www.frontiersin.org/article/10.3389/fpsyt.2019.00810/full

32. Scahill L, Challa SA. Repetitive behavior in children with autism spectrum disorder: Similarities and differences with obsessive-compulsive disorder. In: Mazzone L, Vitiello B, editors. Psychiatric Symptoms and Comorbidities in Autism Spectrum Disorder. Cham: Springer International Publishing; 2016. p. 39–50.

33. Van Ameringen M, Patterson B, Simpson W. DSM-5 obsessive-compulsive and related disorders: Clinical implications of new criteria. Vol. 31, Depression and Anxiety. Blackwell Publishing Inc.; 2014. p. 487–93.

34. Lecavalier L. Behavioral and emotional problems in young people with pervasive developmental disorders: Relative prevalence, effects of subject characteristics, and empirical classification. J Autism Dev Disord [Internet]. 2006 Nov [cited 2020 Feb 26];36(8):1101–14. Available from: http://www.ncbi.nlm.nih.gov/pubmed/16897387

35. Gadow KD, Devincent CJ, Pomeroy J, Azizian A. Comparison of DSM-IV symptoms in elementary school-age children with PDD versus clinic and community samples. Autism [Internet]. 2005 Oct 30 [cited 2019 Apr 15];9(4):392–415. Available from: http://www.ncbi.nlm.nih.gov/pubmed/16155056

36. Wijnhoven LAMW, Creemers DHM, Vermulst AA, Granic I. Prevalence and risk factors of anxiety in a clinical Dutch sample of children with an autism spectrum disorder. Front Psychiatry. 2018 Mar 2;9(MAR).

37. Case R, Hayward S, Lewis M, Hurst P. Toward a neo-Piagetian theory of cognitive and emotional development. Dev Rev [Internet]. 1988 Mar 1 [cited 2019 Jun 17];8(1):1–51. Available from: https://www.sciencedirect.com/science/article/pii/027322978890010X

38. Fischer KW, Shaver PR, Carnochan P. A skill approach to emotional development: From basic-to subordinate-category emotions. -PsycNET. In: W. Damon, editor. The Jossey-Bass social and behavioral science series Child development today and tomorrow [Internet]. San Francisco, CA, US: Jossey-Bass; 1989 [cited 2019 Jul 10]. p. 107–36. Available from: https://psycnet.apa.org/record/1988-98806-006

39. Broeren S, Muris P. The Relation Between Cognitive Development and Anxiety Phenomena in Children. J Child Fam Stud [Internet]. 2009 Dec [cited 2019 Jul 10];18(6):702–9. Available from: http://www.ncbi.nlm.nih.gov/pubmed/19855850

40. Muris P, Merckelbach H, Luijten M. The connection between cognitive development and specific fears and worries in normal children and children with below-average intellectual abilities: a preliminary study. Behav Res Ther [Internet]. 2002 Jan [cited 2019 Jul 10];40(1):37–56. Available from: http://www.ncbi.nlm.nih.gov/pubmed/11762426

41. Vancu G. Anxiety of children with intelectual disability. Agora Psycho-Pragmatica [Internet]. 2018 Nov 29 [cited 2019 Jul 10];12(1):107–14. Available from: https://www.uav.ro/jour/index.php/app/article/view/941

42. Rieske RD, Matson JL, Davis TE. The Moderating Effect of Autism Symptomatology on Anxiety Symptoms. J Dev Phys Disabil [Internet]. 2013 Oct 9 [cited 2019 Jul 10];25(5):517–31. Available from: http://link.springer.com/10.1007/s10882-012-9330-0

43. Deák GO. Interrelations of language and cognitive development. In: Brooks P., Kampe V, editors. Encyclopedia of Language Development [Internet]. NewYork: Sage; 2014 [cited 2020 Feb 6]. Available from: https://www.researchgate.net/publication/265905729_Interrelations_of_language_and_cognitive_development

44. Bernstein GA, Layne AE, Egan EA, Nelson LP. Maternal phobic anxiety and child anxiety. J Anxiety Disord [Internet]. 2005 Jan [cited 2020 Feb 6];19(6):658–72. Available from: https://linkinghub.elsevier.com/retrieve/pii/S0887618504000829

